# Low and high infection dose transmissions of SARS-CoV-2 in the first COVID-19 clusters in Northern Germany

**DOI:** 10.1101/2020.06.11.20127332

**Authors:** Susanne Pfefferle, Thomas Günther, Robin Kobbe, Manja Czech-Sioli, Dominic Nörz, René Santer, Jun Oh, Stefan Kluge, Lisa Oestereich, Kersten Peldschus, Daniela Indenbirken, Jiabin Huang, Adam Grundhoff, Martin Aepfelbacher, Johannes K. Knobloch, Marc Lütgehetmann, Nicole Fischer

## Abstract

**Objectives:** We used viral genomics to deeply analyze the first SARS-CoV-2 infection clusters in the metropolitan region of Hamburg, Germany. Epidemiological analysis and contact tracing together with a thorough investigation of virus variant patterns revealed low and high infection dose transmissions to be involved in transmission events.

**Methods:** Infection control measures were applied to follow up contract tracing. Metagenomic RNA- and SARS-CoV-2 amplicon sequencing was performed from 25 clinical samples for sequence analysis and variant calling.

**Results:** The index patient acquired SARS-CoV-2 in Italy and after his return to Hamburg transmitted it to 2 out of 132 contacts. Virus genomics and variant pattern clearly confirms the initial local cluster. We identify frequent single nucleotide polymorphisms at positions 241, 3037, 14408, 23403 and 28881 previously described in Italian sequences and now considered as one major genotype in Europe. While the index patient showed a single nucleotide polymorphism only one variant was transmitted to the recipients. Different to the initial cluster, we observed in household clusters occurring at the time in Hamburg also intra-host viral species transmission events.

**Conclusions:** SARS-CoV-2 variant tracing highlights both, low infection dose transmissions suggestive of fomites as route of infection in the initial cluster and high and low infection dose transmissions in family clusters indicative of fomites and droplets as infection routes. This suggests (1) single viral particle infection can be sufficient to initiate SARS-CoV-2 infection and (2) household/family members are exposed to high virus loads and therefore have a high risk to acquire SARS-CoV-2.

## Introduction

SARS-CoV-2, first emerged in late 2019 as the alleged cause of a cluster of viral pneumonia cases in Wuhan (1). The virus and its associated disease, COVID-19 have since given rise to a worldwide pandemic. Significant findings addressing epidemiology, containment measures, transmission dynamics and course of the disease have been published worldwide in short intervals. However, questions with regard to the infectivity of the virus, how many viral particles are sufficient for infection, are droplets or fomites the predominant transmission routes, still need to be addressed in more detail.

Viral sequence data were immediately publically available and online tools such as GISAID provided graphical interfaces for phylogenetic network analyses (1–6). Viral genomics can further be used to deepen our understanding of infection routes and viral sequence variability, which might be contributing to the clinical course of the disease.

Previous reports suggest frequent human-to-human transmission together with a wide range of clinical severity, including patients showing mild or no symptoms with at the same time high concentrations of viral RNA detectable in respiratory samples (7–9).

Interestingly, a recent report describes the dynamics of viral shedding and transmissibility of SARS-CoV-2 with more than 44% of transmission occurring prior to symptom onset and viral loads peaking before or at the time of symptom onset (10). This implies that not only health care workers need adequate protection due to more frequent contact with infected patients (with high viral load) but especially household/family members are at high risk due to high exposure and close contact.

While the viral load effect on incidence of infection and severity of the disease is currently discussed, however, infecting dose measurements are highly challenging and can only be adequately addressed in animal models.

We here use viral genomics and variant calling to follow up viral transmission in the initial COVID-19 cluster in Hamburg and other small household clusters occurring at the time in travel returnees in Hamburg. We show here for the first time using viral variant tracing that low infection dose and high infection dose transmissions contribute to the spread of SARS-CoV-2. Overall by analysing 25 clinical samples using viral genomics and variant analysis we find evidence for both, intra-host variant transmission indicative of high viral dose transmissions by droplets and transmission of only one sub-consensus variant indicative of a low dose infection event.

## Methods

### Patients

The index patient is a physician in his 60s with no underlying diseases. 10d past his initial SARS-CoV-2 positive PCR test he was precautionary admitted to a hospital for 4 days where a chest-CT scan showed characteristic ground-glass opacities (supplementary figure S1).

Patient 1, a male laboratory worker in his late 40s, with type II diabetes and high BMI. He developed severe symptoms with dry cough, muscle pain, high fever and acute respiratory distress syndrome (ARDS) with ICU admission and ECMO treatment.

Patient 2, a relative of patient 0 living in the same household, in her 60s, no comorbidities known, developed a mild cough 3d after initial testing. She never developed other symptoms of COVID-19 disease (supplementary Tables S2-3).

Additional 20 oropharyngeal samples were collected between 1^st^ and 8^th^ of March 2020 during SARS-CoV-2 outpatient testing at the University Medical Center Hamburg-Eppendorf, UKE, following the Robert Koch Institute’s, RKI, recommendations at that time (individuals with respiratory symptoms returning from countries/regions defined as high risk regions and/or individuals with contacts to positively tested persons).

The local ethics committee of the City of Hamburg approved the study (PV7306). All studies were carried out in keeping with local legal and regulatory requirements.

### Epidemiological investigation

Contact persons of the index patient and patient 1 were identified by the UKE infection prevention and control team together with public health officials. Classification of contacts is summarized in supplementary Table S1. All employees with any contact were tested for SARS-CoV-2 at the day of confirmed infection of the index patient. Persons with positive SARS-CoV-2 PCR were sent to home quarantine until the virus became undetectable and were closely monitored for symptom history. Personnel with high-risk contacts were sent to home quarantine for 14 days and were tested at the end of their quarantine. Employees with low risk contacts remained on duty and voluntary testing was provided on day 7 after the last contact to the index patient.

### Detection of SARS-CoV-2 RNA via RT-PCR in clinical samples

Oropharyngeal swabs were preserved in modified Amies medium (Copan E-swab, Copan, Italy). Respiratory samples, serum, urine and stool samples were taken without additives, 1:1 mixed with Roche PCR Media kit buffer (Roche, USA). Screening PCR for SARS-CoV-2 RNA was performed as described previously (11, 12).

### Cell culture and virus isolation

Vero E6 cells (ATCC® CRL-1586) were propagated in DMEM containing 3% FCS, 1% Penicillin/ Streptomycin, 1 % L-Glutamine, 1 % Sodium pyruvate, 1 % non-essential amino acids (Gibco/ Thermo Fisher, Waltham, USA) under standard culture conditions. For virus isolation, cells were washed with PBS prior to infection with the native clinical material. After adsorption for 1h at 37°C, fresh DMEM was added. Virus replication was assessed by RT-PCR.

### Immunofluorescence assays

Assays were performed as described before (13, 14).

### ELISA for detection of SARS-CoV-2 IgG

A commercially available ELISA for detection of IgG against SARS-CoV-2 (Euroimmun, Lübeck, Germany) was used on the Euroimmunanalyzer I (Euroimmun, Lübeck, Germany).

### SARS-CoV-2 Amplicon Sequencing

SARS-Cov-2 amplicon sequencing was based on the nCoV-2019 sequencing protocol from the Artic network project (artic.network/ncov-2019) with the following modifications. Reverse transcription was performed in the presence of 2.4µM random primer mix (hexamer and anchored-dT primer (dT23VN)) After RNase H digestion multiplex PCR was performed using the primer scheme version 3 with 196 primers (supplementary Table S4) generating 400bp amplicons. Adaptor ligation for Illumina compatible multiplex sequencing was achieved by using the NEBNext DNA Ultra II Library Prep Kit and the NEBNext Multiplex Oligos (96 Unique Dual Index Primer Pairs; New England Biolabs). Multiplex sequencing was performed on Illumina MiSeq, 500cycle MiSeq v2 kit (Illumina).

### Unbiased metagenomic RNA sequencing

NGS libraries were prepared from each sample using SMARTer Stranded Total RNA-Seq Kit v2 - Pico Input Mammalian (Takara Bio Europe, Saint-Germain-en-Laye, France). Multiplex-sequencing was performed on an Illumina NextSeq, 300 cycles, PE protocol.

The number of reads per sample is summarized in supplementary tables S5-S6. Samples were analyzed using an in-house pathogen detection pipeline (15–18).

### Bioinformatic analysis of metagenomic - and amplicon sequencing

Illumina paired end amplicon sequencing reads were merged using pear (19), filtered by amplicon length and the presence of correct amplicon primer sequences at both ends and aligned to NC_045512.2 using minimap2 (20) with default settings for short read alignment.

Variants were called using freebayes Bayesian haplotype caller v1.3.1 (21) with ploidy and haplotype independent detection parameters to generate frequency-based calls for all variants passing input thresholds (-K -F 0.01). Input thresholds were set to 10 variant supporting reads with a minimum base quality of 30 (-C10 -q30). Only high confidence variants present in > 33% of reads within at least one individual sample were included and annotated using ANNOVAR (22).Variants located in non-coding regions or representing frameshift, stopgain or startloss were deleted. Data were clustered by hclust clustering method ward.D2.

To detect subgenomic mRNAs reads containing 10 upstream bases and the 6 core element bases of the transcription-regulating sequence (TRS-L) fused to the 10 bases following the TRSs of the subgenomic mRNA bodies (TRS-B) of the individual subgenomic mRNAs were counted.

All metagenomic and amplicon based sequences used in this work (after removal of human sequences) are publicly accessible from the European Nucleotide Archive with the study accession number PRJEB38546.

## Results

In February 2020 a physician (index patient) returned from Trentino, Italy, at this time, not defined as a risk area for contracting SARS-CoV-2 (23). 2 days after his return, he developed symptoms of a cold, stopped his duty and stayed in home quarantine together with a relative thereafter. He was laboratory confirmed as SARS-CoV-2 positive (Figure 1).

**Figure 1:**
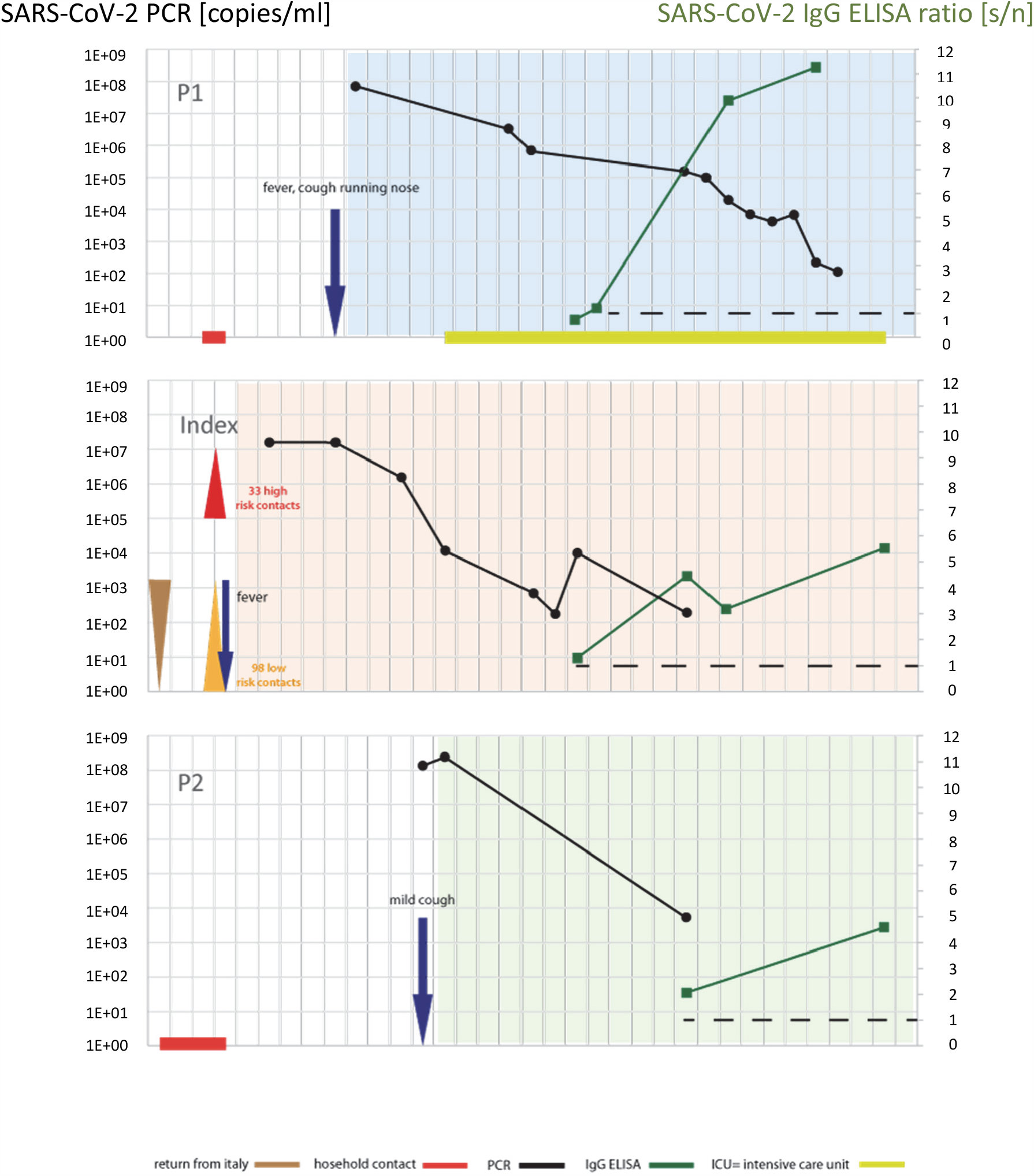
Overview of transmission events and course of the disease in the first COVID-19 cluster in Northern Germany. SARS-CoV-2 RNA loads (black line, left y-axis) in respiratory samples and SARS-CoV-2 IgG ELISA ratios (green line, right y-axis) from the index patient (centered panel), patient 1 (upper panel) and patient 2 (lower panel). Brown triangle indicates initial infection of the index patient in Italy; low and high risk contacts of the index patient are shown as orange and red triangles, respectively. Initial occurrence of symptoms is indicated by an arrow. ICU treatment of patient 1 is labelled with a yellow rectangle. The dashed lines indicate threshold for reporting positive results of the ELISA (= signal to noise ratio, s/n ≥1).

During these 2 days patient 0 had contact with 131 persons at the workplace. Out of these, 33 were high risk (category I) and 98 were low risk contacts (category II). These contacts and a relative of patient 0 were tested negative on February 28th. One high risk workplace contact (patient 1) who had two contacts with Patient 0 developed symptoms 3d later and was subsequently tested positive. Similarly, a relative of patient 0 (patient 2) was tested positive for SARS-CoV-2 at that time. All other high risk contacts, retested at day 7 (n=17) and/or at day 14 (n=28) after the last contact with patient 0 stayed SARS-CoV-2 negative. 40 low risk contacts were tested negative at day 7 after the last contact with patient 0.

### SARS-CoV-2 infection course in the patients belonging to the index cluster

SARS-CoV-2 viral RNA was monitored by RT-PCR in respiratory specimens from patients 0 - 2 at different time points of infection. High viral loads were observed in all three patients early in the infection, independent of clinical symptoms (Figure 1, Tables S2-S3). In patients 0 and 2 the viral loads declined 1000-fold and 200-fold, respectively, within a 5 day period following the first day of positivity (Figure 1). In the critically ill patient 1, however, viral loads declined only 8-fold and remained high over a prolonged time period. Furthermore, while on ECMO treatment, he also showed moderate viral loads in the serum, suggesting a systemic viral infection (Table S3).

Interestingly, from an oropharyngeal swab of patient 2, who stayed asymptomatic, infectious SARS-CoV-2 could be successfully propagated in cell culture (24) suggestive of active viral replication. This is furthermore supported by our data obtained from RNA shotgun sequencing, in which we detect reads spanning the SARS-CoV-2 leader sequence and regions of subgenomic RNAs (sgRNAs) (Table 1). All three patients seroconverted (Figure 1, Table S3), within 10-14 d after the first positive PCR report.

**Table 1:** Subgenomic RNA detection by shotgun RNA-Seq.

| Sample | L<sup>a</sup> | S | ORF3a | E | M | ORF6 | ORF7a | ORF8 | N | Total sg<sup>b</sup> |
| --- | --- | --- | --- | --- | --- | --- | --- | --- | --- | --- |
| P01 | 3 | 0 | 0 | 0 | 0 | 0 | 0 | 0 | 6 | 6 |
| P03 | 0 | 0 | 0 | 0 | 0 | 0 | 0 | 0 | 0 | 0 |
| P02 | 0 | 0 | 0 | 0 | 0 | 0 | 0 | 2 | 3 | 5 |
| P06 | 7 | 0 | 4 | 2 | 2 | 0 | 6 | 0 | 14 | 28 |
| P07 | 1 | 2 | 0 | 0 | 0 | 0 | 0 | 0 | 2 | 4 |
| P05 | 0 | 0 | 0 | 0 | 0 | 0 | 0 | 0 | 0 | 0 |
| P09 | 0 | 0 | 0 | 0 | 0 | 0 | 0 | 0 | 14 | 14 |
| P10 | 0 | 0 | 0 | 0 | 0 | 0 | 0 | 0 | 3 | 3 |
| P11 | 0 | 0 | 0 | 0 | 0 | 0 | 0 | 0 | 3 | 3 |
| P04 | 0 | 0 | 0 | 16 | 0 | 0 | 8 | 0 | 3 | 27 |
| P13 | 2 | 0 | 0 | 0 | 0 | 0 | 0 | 0 | 1 | 1 |
| P15 | 0 | 0 | 0 | 0 | 0 | 0 | 0 | 0 | 0 | 0 |
| P16 | 0 | 0 | 0 | 0 | 0 | 0 | 1 | 0 | 2 | 3 |
| P18 | 0 | 0 | 0 | 0 | 0 | 0 | 0 | 0 | 15 | 15 |
| P21 | 7 | 2 | 0 | 0 | 0 | 0 | 0 | 0 | 13 | 15 |
| P22 | 11 | 0 | 0 | 0 | 0 | 1 | 1 | 0 | 13 | 15 |
| P25 | 0 | 0 | 0 | 0 | 0 | 0 | 0 | 0 | 0 | 0 |
| P30 | 0 | 0 | 0 | 0 | 0 | 0 | 0 | 0 | 0 | 0 |
| P33 | 2 | 0 | 10 | 0 | 2 | 0 | 0 | 1 | 59 | 72 |
| P35 | 0 | 0 | 0 | 0 | 5 | 0 | 0 | 0 | 1 | 6 |
| P13 | 3 | 0 | 0 | 0 | 1 | 0 | 0 | 0 | 0 | 1 |
| P39 | 4 | 0 | 0 | 0 | 0 | 0 | 1 | 0 | 20 | 21 |
| P40 | 1 | 2 | 0 | 0 | 0 | 0 | 0 | 0 | 0 | 2 |
| P48 | 8 | 0 | 0 | 0 | 0 | 0 | 1 | 0 | 5 | 6 |
| P35 | 0 | 0 | 0 | 0 | 0 | 0 | 0 | 0 | 0 | 0 |
| cell culture | 340 | 24 | 6 | 10 | 13 | 24 | 18 | 8 | 87 | 190 |
| total<sup>c</sup> | 389 | 30 | 20 | 28 | 23 | 25 | 36 | 11 | 264 |  |

Summary of subgenomic mRNA-detecting reads spanning the leader sequence fused to the genome region encoding for structural and accessory proteins.
<sup>a</sup> leader sequence within the genomic (+) mRNA <sup>b</sup> total number of reads indicating presence of all subgenomic mRNAs within each sample <sup>c</sup> total number of fragments indicating subgenomic mRNAs across all samples.

### SARS-CoV-2 variant calling allows geographic viral origin analysis and contact tracing

Since we did not obtain high virus coverage by shotgun sequencing over the complete SARS-CoV-2 genome in all samples, we performed SARS-CoV-2 amplicon sequencing. This allowed us to thoroughly characterize viral genomes, confirm the relationship of the viral sequences and identify possible sequence variants. We obtained coverage of more than 98% of all SARS-CoV-2 genomes included (n=25) with 23 samples showing >99.5% coverage, the maximum coverage of amplicon sequencing, due to primer positioning at the very 5’ and 3’ ends. We identified 37 sequence variants, with 26 variants not used for clade or genotype assignment and 9 variants not represented in the GISAID database (Figure 2). Overall, mutations within the nucleotide sequence were relatively rare. We find between 2 to 12 consensus level variants per sample and only few, 1-2, sub-consensus variants occurring in 30% of the samples. Based on the observed SNPs we can clearly cluster the sequences in distinct variant patterns, pattern I, II (II.1-II.4) and III (Figure 2). The clustering is driven by the presence of few SNPs, frequently found in SARS-CoV-2 sequences. These were described e.g. in the leader sequence, position 241 co-occurring with mutations within the nsp3 (nt 3037), RNApol (nt 14408) and spike protein (nt 23403). These co-occurring mutations have been recently described as one major SARS-CoV-2 variant occurring in Europe and suggested to be defined as genotype II (6). The earliest representative of this prevalent SNP pattern in GISAID (3, 25) is in an Italian sequence (Italy/CDG1/2020/412973) entered on February 20^th^. We identified these mutations in one cluster of related infections from persons returning from a weekend of clubbing in Berlin (Figure 2, pattern II.1). In addition to these 4 core variants additional frequent variants are known and here described as sub-clusters of cluster II. Cluster II.2 is defined by an additional frequent mutation of three consecutive bases in the nucleocapsid phosphoprotein, nt 28,881 resulting in two aa changes. This SNP is described in approximately 25% of sequences in GISAID with its first description on February 24^th^, Netherlands/Berlicum_1363564/2020. Sequences isolated from our index patient, patient 1 and 2 are clustering in pattern II.2. The sequences are highly related defined by an additional SNP at position 160, a variant not described in any sequence currently deposited in any database (3, 25). Thus, the mutation at position 160 in combination with the anamnestic data and the chronological development of test positivity in the three patients, proves that patient 1 and -2 acquired SARS-CoV-2 from patient 0.

**Figure 2:**
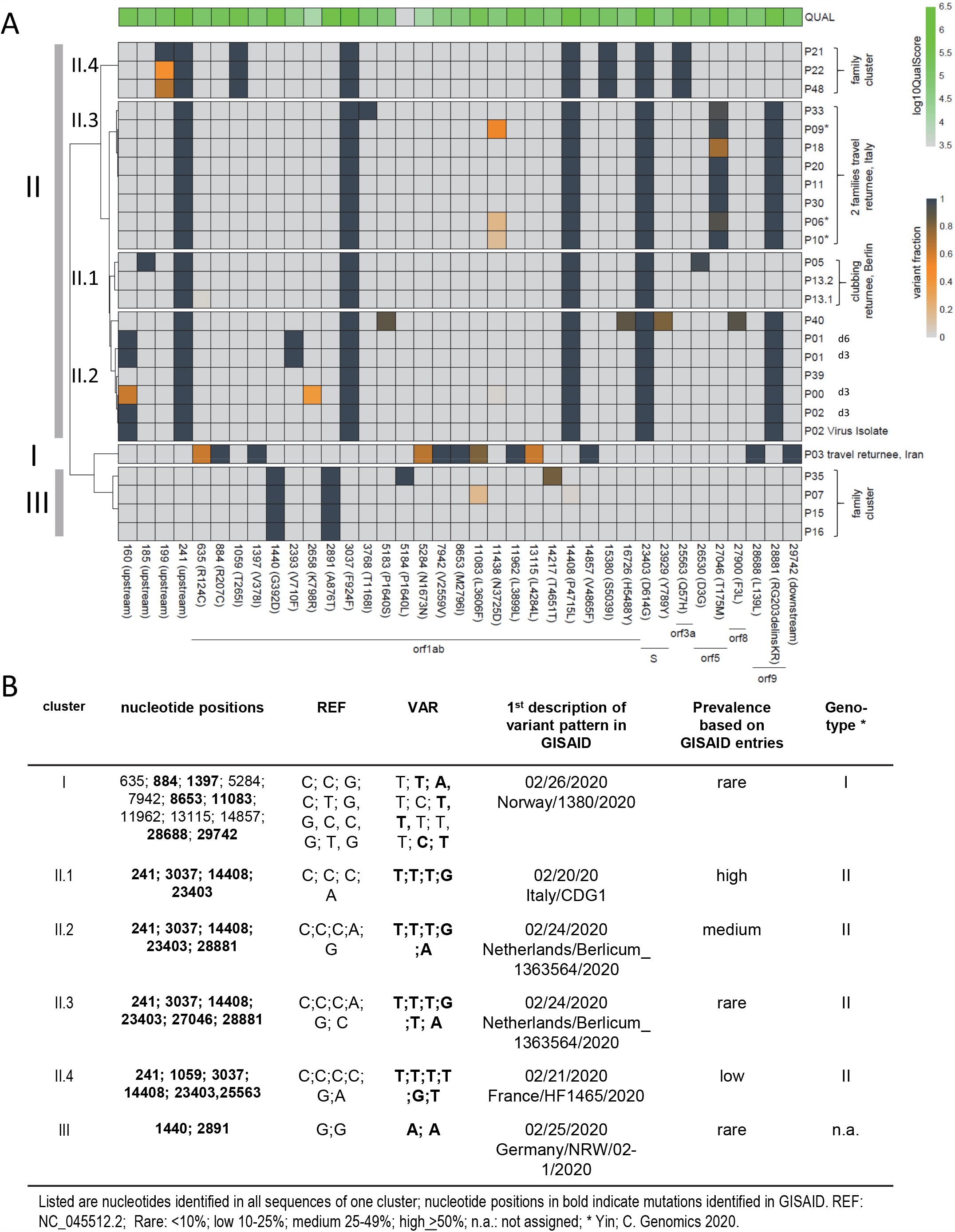
**(A)** Clustering of viral variants of SARS-CoV-2 sequences recovered from the index patient, patient 1, patient 2 and 19 SARS-CoV-2 sequences from respiratory swabs collected in the same time period in comparison to the reference sequence, NC_045521. Nucleotide positions are indicated at the bottom. Included are only variants with sufficient coverage (>10) and SNP being present in more than 33% of all reads in at least one sample. I-III summarizes sequence patterns as defined by SNPs. The frequency of variants is indicated by the heat map ranging from grey (reference), yellow to dark blue (variant). The quality score per individual site is indicated at the top. * indicates members within one family. **(B)** Overview of individual SNPs defining pattern I, II (II.1-II.4) and III with sequence variation in comparison to the reference sequence.

Interestingly, we identify a unique mutation acquired in patient 1, nt 2393 within orf1ab resulting in an aa substitution Val to Phe at position 710 in the non-structural protein nsp2 (Table 2). The nsp2 protein shows relatively little conservation across coronaviruses and is dispensable for replication *in vitro* (26). However, recent reports suggest that nsp2 can modulate the host cell environment, including transcriptional processes (27).

**Table 2:** Nucleotide variants of SARS-CoV-2 sequences in comparison to the reference sequence NC_045512.2.

|  | Index Patient | Patient 1 |  | Patient 2 |
| --- | --- | --- | --- | --- |
| Position [nt] (orf) | d2* | d2* | d9* | d3* |
| 160 (noncoding) | G/A | G/A | G/A | G/A |
| 241 (noncoding) | C/T | C/T | C/T | C/T |
| **2393 (orf1ab, Nsp2)** | **G** | **G/T (V710F)** | **G/T (V710F)** | **G** |
| 3037 (orf1a, Nsp3) | C/T (F924F) | C/T (F924F) | C/T (F924F) | C/T (F924F) |
| 14408 (orf1ab) | C/T (P4715L) | C/T (P4715L) | C/T (P4715L) | C/T (P4715L) |
| 23403 (S) | A/G D614G | A/G D614G | A/G D614G | A/G D614G |
| 28881(orf9, N) | GGG/AAC<br>(RG203/4KR) | GGG/AAC<br>(RG203/4KR) | GGG/AAC<br>(RG203/4KR) | GGG/AAC<br>(RG203/4KR) |

\*time point after SARS-CoV-2 confirmation by PCR.

Cluster II.3 separates from cluster II.2 with the presence of one additional SNP at nt27046 within the coding region of the membrane glycoprotein. Pattern II.4 is mainly defined by the SNPs at positions 241, 1059, 3037, 14408, 15380, 23403 and 25563. It is less frequent and has been first reported in a sequence from France (France/HF1465/2020) on February 21^st^. Cluster III (rarely identified) is characterized by two SNPs at positions 1440 and 2891. It was first reported in a sequence from Germany (Germany/NRW/02-1/2020) on February 25^th^. Finally, pattern I is clearly distinct; this sequence shows 12 SNPs with six of them being previously, February 26^th^, reported in a sequence identified in Norway (Norway/1380/2020), Figure 2B. The SNP at nt 11962 has been rarely reported with its first description in the sequence Wuhan/HBCDC-HB-05/20/20 on January 18^th^. It was recently suggested as being indicative of genotype I (6).

### Sub-consensus SARS CoV-2 variant calling reveals intra-host transmissions

We used paired end amplicon sequencing data for variant calling and to define minority sequences variants, so called sub-consensus viral populations. Of the 37 nucleotide positions with SNPs, we find at 14 positions sub-consensus variants: leader region (nts 160 and 199), leader protein (nt 635), nsp2 (nt, 2658), nsp3 (nts 5183 and 5284), nsp6 (nts 11083, 11438), nsp10 (nt 13115), RNA polymerase (nt 14217), RNA helicase (nt 16726), S (nt 23939), M (nt 27046) and TM (nt 27900). Most of these variants result in non-synonymous mutations with the exception of the variants at nts 5284, 13115 and 14217 causing synonymous mutations.

Interestingly, we identified three sequence clusters in which we can follow transmission events based on the presence/absence of minority sequences at specific nucleotide positions. Within the first cluster we detected patient 0 carrying a sub-consensus variation at position 160 in the leader region. The two recipients, patient 1 and 2, both carry only one variant at position 160, in both cases G (Var) instead of A (Ref). Considering the index patient showing a variant fraction of 0.5 at position 160 this is highly indicative of low dose viral infection in both transmission events. Similarly, in another family cluster (cluster II.4), patients P21, P22 and P48, we observed only one position with multiple variants, position 199. Observed variant fractions are highly suggestive of a low viral dose exposure from P22 or P48 to P21. Interestingly, we observed the transmission of intra-host variants at position 199 from patient P22 to P48 (or vice versa) being indicative of high viral load exposure and transmission. Furthermore, we observed the transmission of intra-host variants in a second household cluster, two families returning from a joint vacation. P06, P09 and P10, which are members of one family, are showing intra-host variants at position 11438 indicative of high infection dose exposure being involved in viral transmission. While P09 shows a variant fraction of 0.5 at position 11438, P06 and P10 both have a variant fraction of 0.2. While we cannot make any conclusions on the order of the infection events, however, the presence of identical minority sequence variants in these three patients is highly indicative of high-infection dose transmission between these family members or between the initial sources of the infection and all three family members. Overall, the presence of identical minority sequence is highly suggestive of high viral load exposure occurring in household/family environment.

### No clinically relevant viral or bacterial co-infections in respiratory samples

We performed unbiased metagenomic RNA sequencing from respiratory specimens of the 25 patients included to detect putative viral and bacterial co-infections (Tables 3, S4-S6).

**Table 3:**
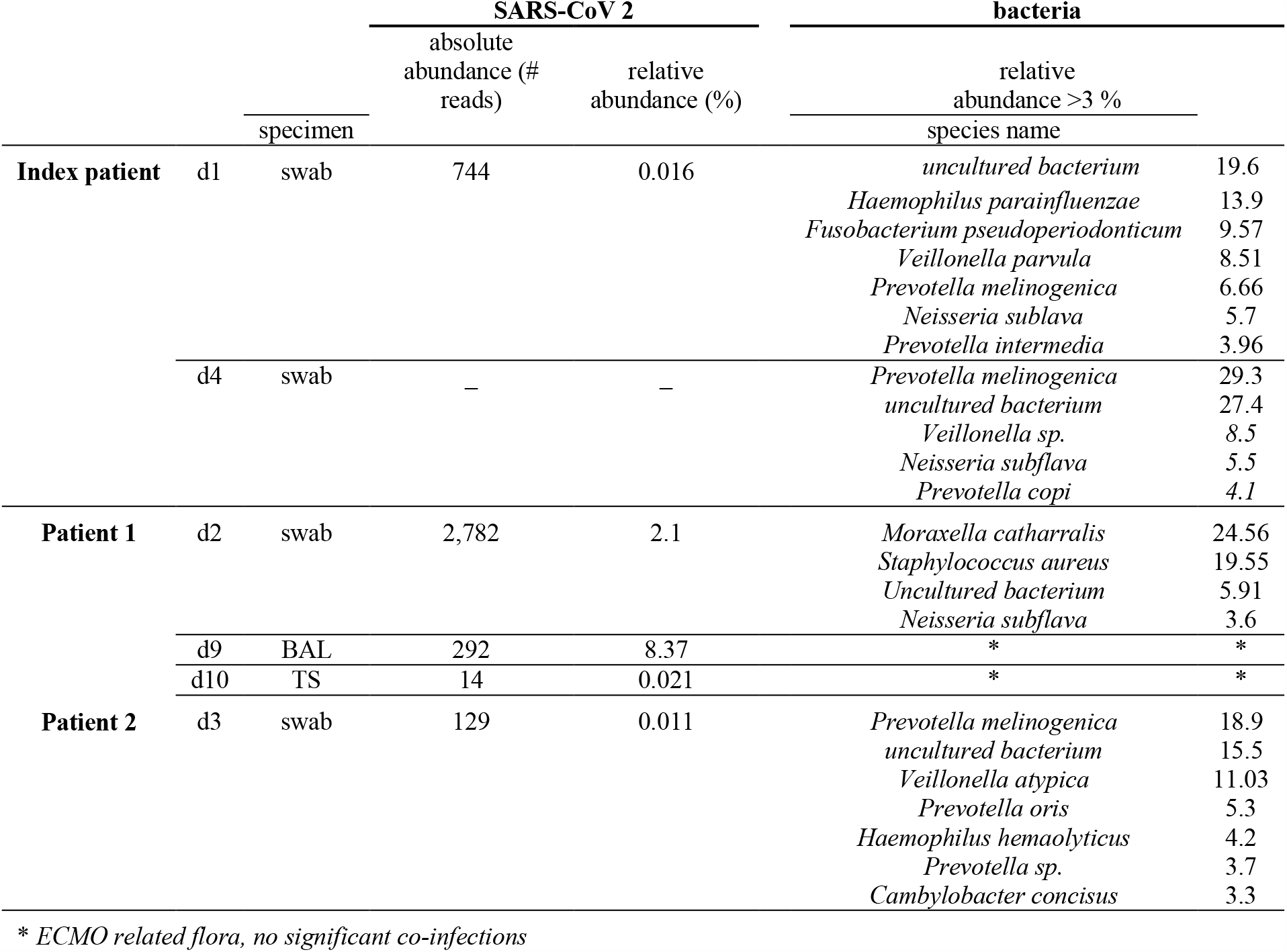
Unbiased metagenomic RNA sequencing describing possible co-infections.

Besides SARS-CoV-2 sequences, no other viral sequences known to be associated with respiratory infections were identified. Furthermore, the identified bacterial sequences in all cases clearly reflected a commensal bacterial flora typical for the oropharyngeal tract. We closely monitored the samples of the index cluster in which we sampled longitudinal respiratory samples of the course of the infection. While we did not observe significant overrepresentation of sequences indicative of bacterial co-infections in the patients 0 or 2, an early throat swap sample from patient 1 exhibited a high proportion of sequences assigned to *Moraxella catharralis* and *Staphylococcus aureus* (24.56% and 19.55% relative abundance, respectively). However, tracheal secretion and BAL fluid from patient 1 at later time points (under ICU treatment) did not find evidence for persistent bacterial co-infection with *Moraxella catharrhalis* or any other pathogen known as a cause for respiratory infection (Table 3).

## Discussion

This work describes the first SARS-CoV-2 positive patient in Northern Germany, documents the tracing of all its contacts and describes the subsequent infection cluster. Thorough epidemiological, clinical, virological and metagenomic analyses of the patients in this first cluster together with 20 SARS-CoV-2 samples from family and household clusters occurring subsequently in the same geographic area, revealed a number of important insights into transmission, clinical course and molecular epidemiology of SARS-CoV-2/COVID-19.

First, based on comparative SNP analysis using 20 SARS-CoV-2 sequences from our study and sequences available in the GISAID database, we identify different virus variant patterns, which allow us to conclude on transmission routes. Transmission of SARS-CoV-2 from the index patient occurred only to two out of 132 contact persons with whom prolonged and unprotected interaction took place. Although we detected mutations between different patient’s samples the overall frequency was relatively low. Our observation is different to a recent report in which high frequency of SARS-CoV-2 mutations was described (28). Differences can be explained by the relatively conservative variant calling approach in our analysis with variant frequency >33% being reported compared to 5% variant frequency as cut-off in the recently published report. Similar to previous reports, we did not find any mutation hot-spot genes in our analysis.

Within the initial cluster (patient 0, 1 and 2), we identify a single non-synonymous mutation gained in the prevalent virus sequence recovered from patient 1. This mutation has not been described before and results in an aa substitution at position 710 of the non-structural protein nsp2. While we have no indication that this aa substitution has functional consequences for viral replication or host interaction, it is interesting that nsp2 is dispensable *in vitro* and not very well conserved between coronaviruses. Interestingly, mutations within nsp2 have been described (26, 29, 30) suggesting a more stable nsp2 protein and thereby contributing to higher pathogenicity (29).

Second, to our knowledge we describe the first report of intra-host variant transmissions of SARS-CoV-2 in patients. We report low dose and high dose transmission of SARS-CoV-2 occurring in family/household clusters. In our initial cluster, we observe only low dose viral transmission occurring at the workplace and in the household, respectively. However, in two family clusters in the early epidemic in Hamburg, we observe transmissions of minority sequence variants between household members indicative of high viral load/droplet exposure. While we cannot draw any conclusion on infection dose and disease outcome (e.g. since in the initial cluster the severity of clinical symptoms was independent of viral loads early in infection), our study shows for the first time that in settings of household clusters transmission of minority virus variants occur, which is indicative of high infection dose transmissions by droplets. Our data suggest that (1) low infection dose is sufficient for establishing a SARS-CoV-2 infection and (2) close contacts, household members are at high risk to SARS-CoV-2 infection similar to healthcare workers if not taking appropriate protection measurements.

## Data Availability

All data generated or analysed during this study are included in this published article.

## Acknowledgement

We are grateful to all staff members at the University Medical Center Hamburg-Eppendorf supporting the diagnosis and management of COVID-19 patients. We thank Svenja Reucher and Kerstin Reumann for excellent technical support and Uwe Ganz for graphical illustration. We gratefully acknowledge the authors, originating and submitting laboratories of the sequences from GISAID’s EpiFlu™ Database1 on which part of this research is based.

## Author information

SP, RK, TG, AG, MA, JK, ML and NF designed the study. SP, TG, MC, DI, DN, NF performed literature search; SP, RK, TG, RS, MA, JK, ML and NF wrote the manuscript; SP, RK, TG, MC, DN, JO, SK, LO, KP, DI, JK, ML and NF collected the data. TG, AG and JH performed the bioinformatics analysis; SP, TG, MC, DI, ML and NF generated the figures and tables.

## Declaration of interest

We declare no competing of interest.

## Supplementary Data

Supplementary Figure S1: CT scan of the index patient

Supplementary Table S1: Category of contact to confirm COVID-19 cases

Supplementary Table S2: Clinical parameters obtained in longitudinal samples of P0 and P2.

Supplementary Table S3: Results obtained by microbiological diagnostics.

Supplementary Table S4: Primers used in multiplex PCR for Amplicon Sequencing

Supplementary Table S5: Summary of next generation sequencing data, P0, P1 and P2.

Supplementary Table S6: Viral loads and number of SARS-CoV-2 mapped reads and viral genome coverage.

Supplementary Table S7: Summary of unbiased metagenomic RNA sequencing data.

## Notes

### Competing Interest Statement

The authors have declared no competing interest.

### Clinical Trial

does not apply

### Funding Statement

no funding

### Author Declarations

The local ethics committee of the City of Hamburg approved the study (PV7306).

